# Tanscatheter aortic valve replacement for asymptomatic aortic stenosis - A revisited and contrarian meta-analysis

**DOI:** 10.1101/2025.03.11.25323785

**Authors:** James M. Brophy

## Abstract

**Importance:** Using aggregated data, two recent meta-analyses have concluded that early aortic valve replacement (AVR) was associated with reduced adverse events compared to clinical surveillance in severe but asymptomatic aortic stenosis. However, individual patient data was not used and the possibility and extent of bias due to the unblinding trial design were not considered.

**Objective:** Using reconstructed individual patient level data, the possiblity of early bias was investigated and a meta-analysis of longer term benefits was performed using one year landmark data.

**Evidence Review:** Four randomized trials, as identified from previous systematic reviews, showed important clinical and statistical heterogeneity in year one AVR crossovers to cardiovascular hospitalizations. To minimize any early bias from unblinding, one year landmark analyses were performed separately for each trial and combined in a Bayesian (hierarchical) meta-analysis.

**Findings:** The largest trial with a TAVR intervention arm was the only trial to show improved outcomes in the first year, driven almost completely by an approximate two fold increase in the crossover rate compared to previous SAVR intervention trials. A one year landmark meta-analysis showed no long term benefit for AVR compared to CS for the primary outcome of mortality and cardiovascular hospitalizations for any individual study or for the pooled result (RR 0.70, 95% CI 0.34 - 1.08).

**Conclusions and Relevance:** The early benefit with TAVR in asymptomatic patients with severe aortic stenosis appears more driven by bias than by efficacy. Landmark analysis accounting for this potential bias show no longer term advantage for early AVR compared to clinical surveillance in this population.

**Key Points:** *Question:* Does early intervention for severe asymptomatic aortic stenosis improve patient outcomes compared to clinical surveillance.?

*Findings:* A systematic review suggested early benefits were likely attributable not to interventional efficacy but rather bias due to an unblinded design for a subjective outcome. A one-year landmark meta-analysis showed no long term benefit for early intervention compared to clinical surveillance for the primary composite outcome of mortality and cardiovascular hospitalizations (RR 0.70, 95% CI 0.34 - 1.08).

*Meaning:* After accounting for possible early bias, landmark meta-analysis shows no longer term advantage for early intervention compared to clinical surveillance in this population.

## Introduction

Surgical aortic valve replacement (SAVR) or transcatheter aortic vale replacement (TAVR) are established treaments for patients with symptomatic aortic stenosis (AS). Following a natural history study of AS(1), it was concluded 35 years ago that “operative treatment is the most common cause of sudden death in asymptomatic patients with AS” and consequently it has been standard practice to wait for the appearance of symptoms before proposing an intervention. However, a recent viewpoint(2), co-authored one of the previous authors(1) concluded “… the time has come to recommend AVR for asymptomatic patients with severe AS”. In support of this evolving viewpoint, four recent randomized clinical trials[(3)](4)[(5)](6) and a meta-analysis(7) were cited. A second recent meta-analysis(8) has also concluded that that early AVR provides clinical benefits by reducing adverse events in asymptomatic severe AS patients and that clinical guidelines should be revised accordingly. As this would represent a major shift in clinical practice, a detailed re-examination of the supporting evidence seems appropriate.

## Methods

Four RCTs[(3)](4)[(5)](6) were concordantly identified from two recent systematic reviews and meta-analyses[(7)](8) of aortic valve replacement (AVR) versus clinical surveillance (CS) in asymptomatic patients with severe aortic stenosis. The trial characteristics and outcome data analyzed herein were extracted directly from the original publications. The pooled average CS crossover to AVR data from the three trials with predominately surgical intervention arms were modeled with a Bayesian hierarchical logit-link model and vague priors and compared to the crossover rate from the EARLY_TAVR intervention trial.

The meta-analysis of the outcome data employed individual patient data (IPD) in contrast to the previous meta-analyses[(7)](8) which used aggregate data. The IPD approach was complicated by the refusal of individual trialists to share their data. Notwithstanding this impediment, IPD can be approximated by extracting the data from the published cumulative incidence curves(9). The aggregate cumulative incidence data were, key time points and numbers at risk, were extracted for each study group and then transformed into a format suitable for survival analysis. Through interpolating between cumulative incidence values and calculating changes in risk, events and censorings between the time points were estimated. The simulated IPD therefore mimics the original datasets and allow replication of the published Cox proportional hazards models. Additionally, using this reconstructed patient-level survival data from the published aggregated summaries allowed the generation of Kaplan-Meier survival curves and the performance of landmark analyses.

The IPD approach, with the creation of a 1 year landmark dataset, provided a workaround to any potential early bias due to unblinded trial design. A meta-analysis of the landmark dataset employed a hierarchical Bayesian exponential survival model. Each study, *s*, has a study-specific baseline log-hazard (*αs*) drawn from a Normal(*μ*_*α*_, *σ*_*α*_)prior. A global effect parameter *β*(the log hazard ratio for the intervention) is also estimated. Vague Priors were assumed:

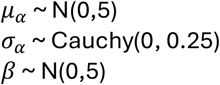

Each individual, *i*, is assumed to have a constant hazard rate

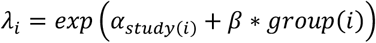

where *α*_*study*(*i*)_ is the baseline log-hazard specific to the individual’s study, and *β*is the log hazard ratio associated with being in the intervention group.

While the pooled estimate and associated credible interval reflects the uncertainty around the average overall effect, prediction intervals are also highly recommended[(10)](11) as they incorporate the between-study variability and capture the range of plausible effects to be expected in a new or future study, given the current evidence. Because a new study can deviate from the mean effect more than the “average” study (especially if heterogeneity is large), the prediction interval is typically wider than the interval around the pooled estimate.

Analyses were performed using the R(12) ecosystem and the statistical code is available <u>here</u>. The Bayesian hierarchical model was compiled and sampled via cmdstanr(13) with four chains, each running for 1000 warmup and 1000 sampling iterations. Convergence diagnostics were examined, and posterior summaries obtained.

## Results

### Data extraction

The four RCTs that have investigated the role of early intervention compared to clinical surveillance are shown in Table 1. Each study examined a population with severe AS, performed baseline exercise stress testing to assure asymptomatic status, and had randomized treatment and clinical surveillance (CS) arms. Nevertheless, there are major differences between the studies, including the time frames when the studies were performed, the duration of follow-up, choice of the the active treatment arm (one TAVR(3) and three predominately SAVR[(4)](5)(6)), crossover rates, choice of primary endpoints, as well as the intensity and type of follow-up to detect the endpoints.

**Table 1:**
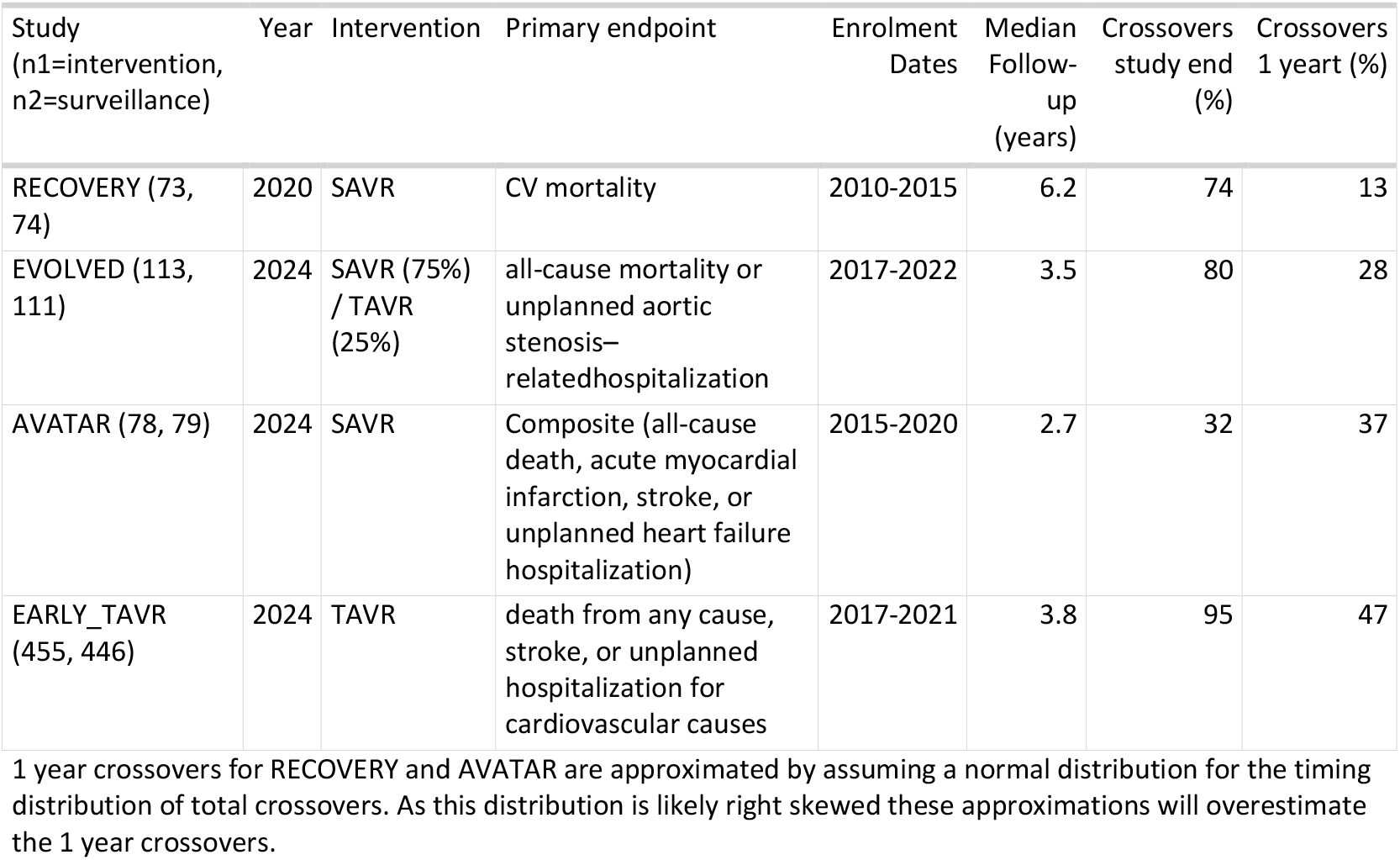
Summary of Randomized Clinical Trials. Key summary data data from from early AS intervention trials.

Table 2 shows the primary outcome for each of the four trials. Given EARLY_TAVR’s(3) large size, almost twice the combined size of the other RCTs[(4)](5)(6), its more contemporary nature, and its exclusive use of TAVR in North American settings, particular attention is directed to this trial.

**Table 2:** Outcomes of Randomized Clinical Trials. Primary ann secondardy outcomes from early AS intervention trials.

| Study<br>(n1=intervention,<br>n2=surveillance) | Primary<br>endpoints<br>(intervention) | Primary<br>endpoints<br>(surveillance) | Mortality<br>(intervention) | Mortality<br>(surveillance) | Hospitalizations<br>(intervention) | Hospitalizations<br>(surveillance) | Stroke<br>(intervention) | Stroke<br>(surveillance) |
| --- | --- | --- | --- | --- | --- | --- | --- | --- |
| RECOVERY<br>(73, 74) | 1 | 11 | 1 | 11 | 0 | 8 | 1 | 3 |
| EVOLVED<br>(113, 111) | 20 | 25 | 16 | 14 | 7 | 19 | 8 | 14 |
| AVATAR (78,<br>79) | 18 | 37 | 13 | 27 | 3 | 13 | 4 | 4 |
| EARLY_TAVR<br>(455, 446) | 122 | 202 | 38 | 41 | 95 | 186 | 19 | 30 |

Figure 1 shows the reconstructed cumulative incidence rates for the four trials and visually well approximate the published figures[(3)](4)[(5)](6). Notwithstanding the differences in the primary endpoints, the most remarkable difference between these four figures, is the extraordinary high one year rate of the primary composite outcome (mortality and unplanned cardiovascular hospitaliztions) in the CS arm of EARLY_TAVR(3), principally driven by TAVR crossover hospitalizations. Remembering that patients in all trials underwent baseline exercise testing to confirm their asymptomatic status, it is remarkable that the EARLY_TAVR(3) one year CS event rate was between 4 and 7 times the previous rates, where the option for leaving the CS group was SAVR. Thus EARLY_TAVR(3) showed a very early and pronounced decrease in the primary outcome in the intervention early group at one year, that is not evident in the other trials, and which dissipated after year one (Figure 2).

**Figure 1.**
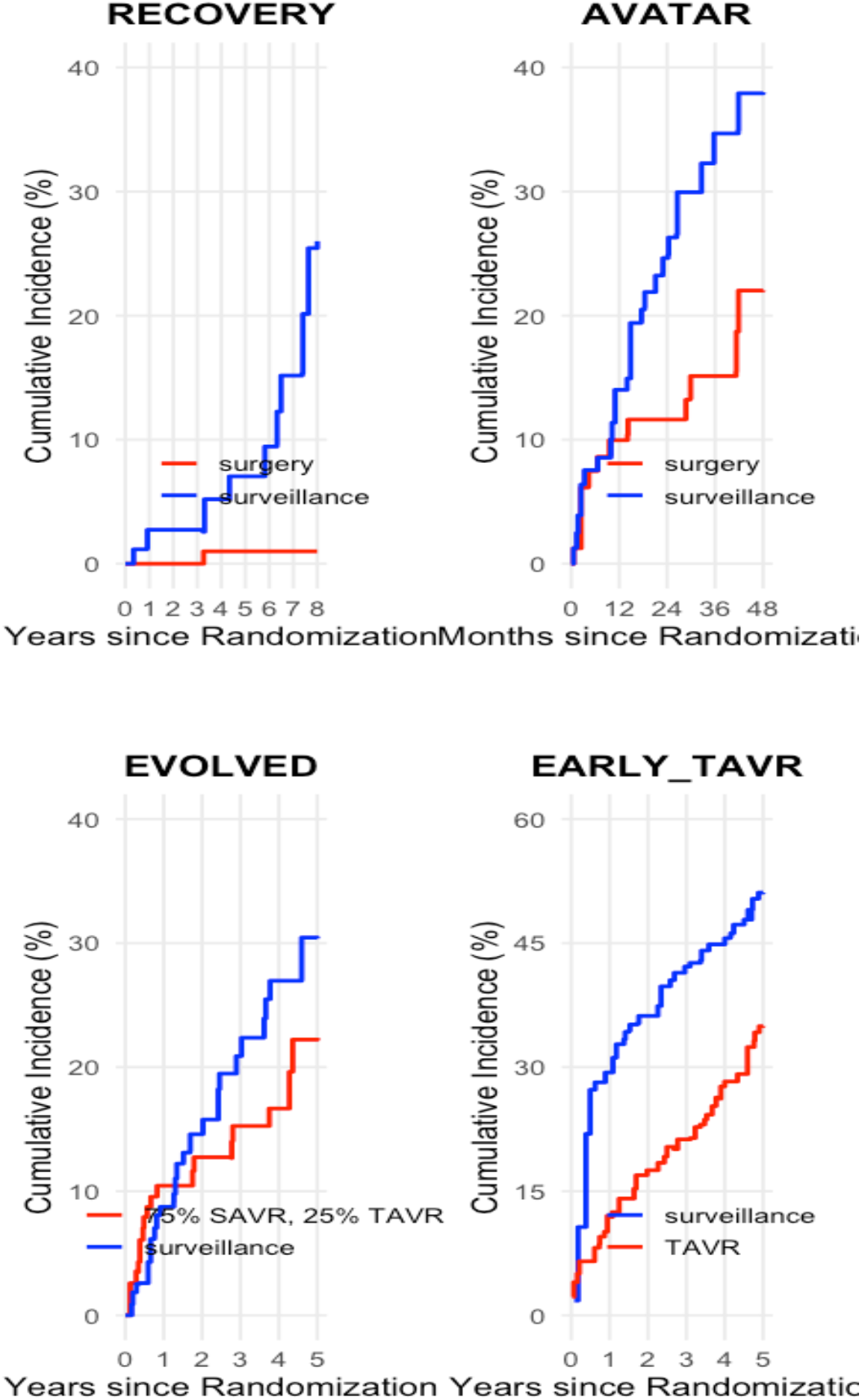
Cumulative incidence plots of 4 RCTs (based on simulated data)

**Figure 2.**
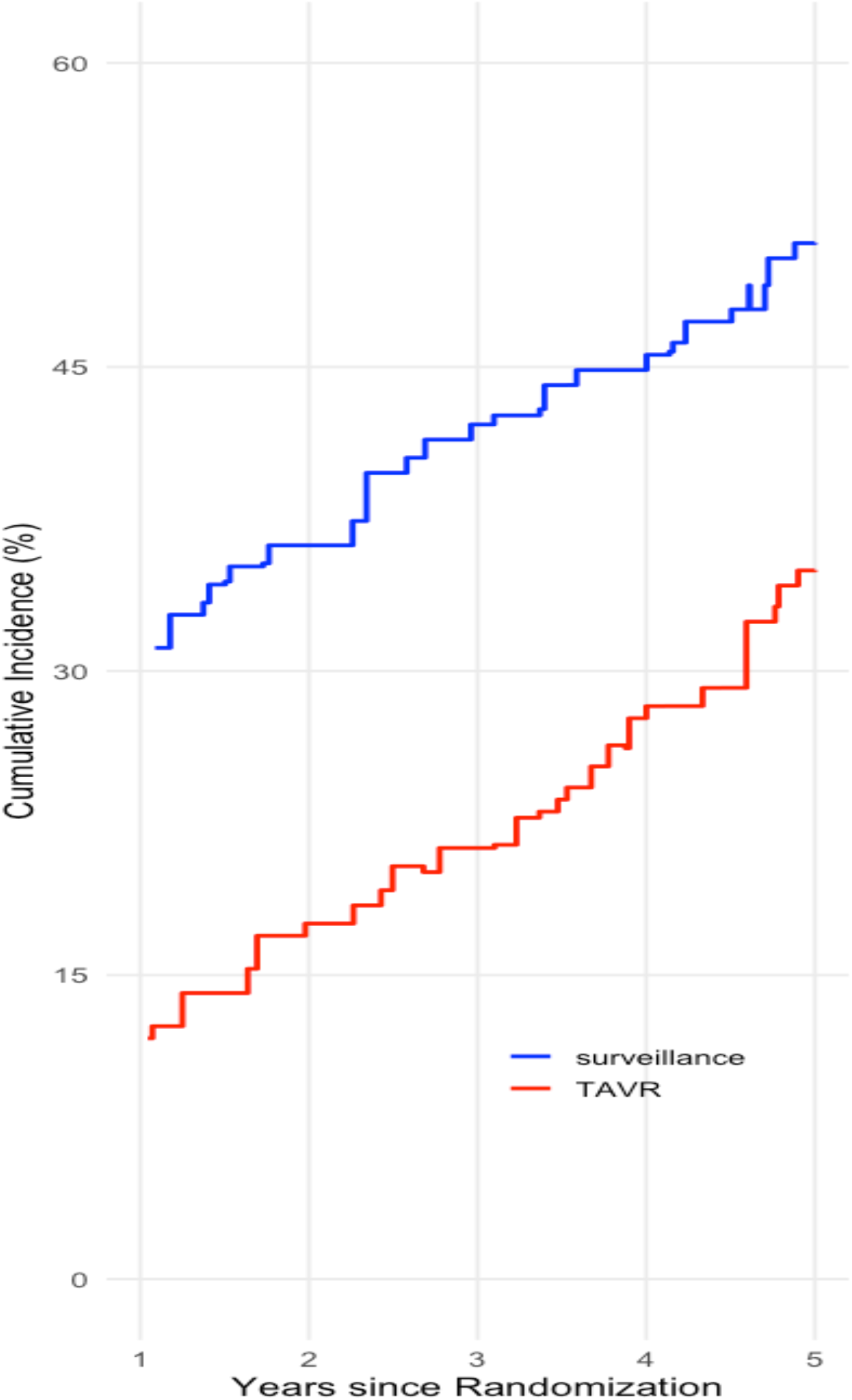
EARLY TAVR Landmark analysis from 1 year

An accuracy test of the generated IPD datasets and the reconstructed cumulative incidence figures involves comparing the calculated and published hazard ratios. Table 3 shows while the published and the calculated hazard ratios are not exactly equal they do generally compare reasonably well for all four trials. An advantage of using simulated IPD, as opposed to aggregqate data, is the ability to perform additional time dependent analyses. For example, the early benefits of intervention in year one can be calculated for all trials. While EARLY_TAVR(3) exhibited a highly statistically signicant decrease in outcomes at 1 year (HR 0.35 (0.26, 0.48: P <0.0001), this was not observed in the other three trials (Table 3). Despite this early benefit in EARLY_TAVR(3), a landmark analysis from 1 year, as predicted by the nearly parallel cuves in Figure 2, showed no statistically significant benefit for TAVR (HR 0.81, 95%CI 0.61, 1.07; P = 0.14) at later time periods.

**Table 3:** Published and calculated HR (95% CI, P value) For primary outcome from early AS intervention trials.

| Study (n1=intervention, n2=surveillance) | Published HR (95% CI, P value) | Calculated 1 Yr HR (95% CI, P value) | Calculated Landmark 1 Yr HR (95% CI, P value) |
| --- | --- | --- | --- |
| RECOVERY, HR 0.09 (0.01,0.67; P = 0.003) | 0.05, (0.01, 0.40 P = 0.004) | NA | 0.06 (0.01, 0.46; P = 0.007) |
| EVOLVED, HR 0.79 (0.44, 1.43; P = 0.44) | 0.71 (0.43, 1.18; P = 0.19) | 1.20 (0.52, 2.78; P = 0.67) | 0.52 (0.27, 1.00; P = 0.051) |
| AVATAR, HR 0.46 (0.23, 0.90; P = 0.02) | 0.51 ( 0.28, 0.92; P = 0.03) | 0.50 (0.22, 1.12; P = 0.09) | 0.52 (0.21, 1.26; P = 0.15) |
| EARLY_TAVR, HR 0.50 (0.40 to 0.63; P < 0.001) | 0.59, (0.46, 0.76; P < 0.001) | 0.35 (0.26, 0.48: P <0.0001) | 0.81 (0.61, 1.07; P = 0.14) |

HR = hazard ratio, CI = confidence interval, NA = not available due to insufficient data

### Bayesian hierarchical models

Given the observed differences in early outcomes between EARLY_TAVR and the other trials, attention was directed to the year one crosssover rates in the CS as each crossover generates a primary outcome via a hospitalization. As shown in Table 1 and Figure 3a year one crossover rates were markedly higher in EARLY_TAVR compared to the pooled average of the other three trials (47% versus 26%) and the probability of these differences exceeding different thresholds is plotted in Figure 3b. For example, there is a 95% probability that the one year difference in CS crossovers between EARLY_TAVR and the other trials is an absolute difference of at least 15%.

**Figure 3a.**
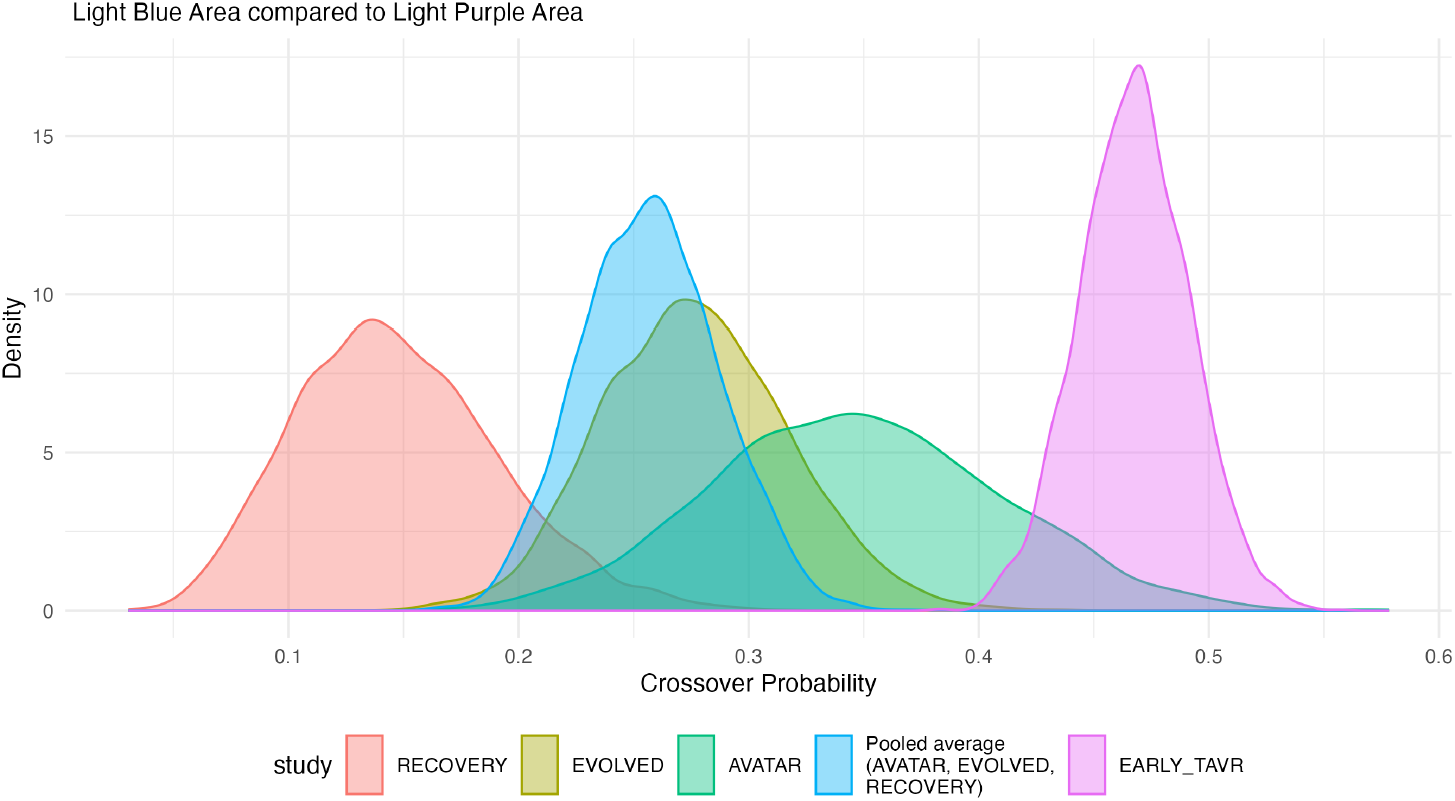
Posterior Distributions of 1 Year Crossover Rates Individual study estimates with pooled average from Bayesian hierarchical model for AVATAR, EVOLVED, and RECOVERY (surgical intervention arms) compared to TAVR intervention trial

**Figure 3b.**
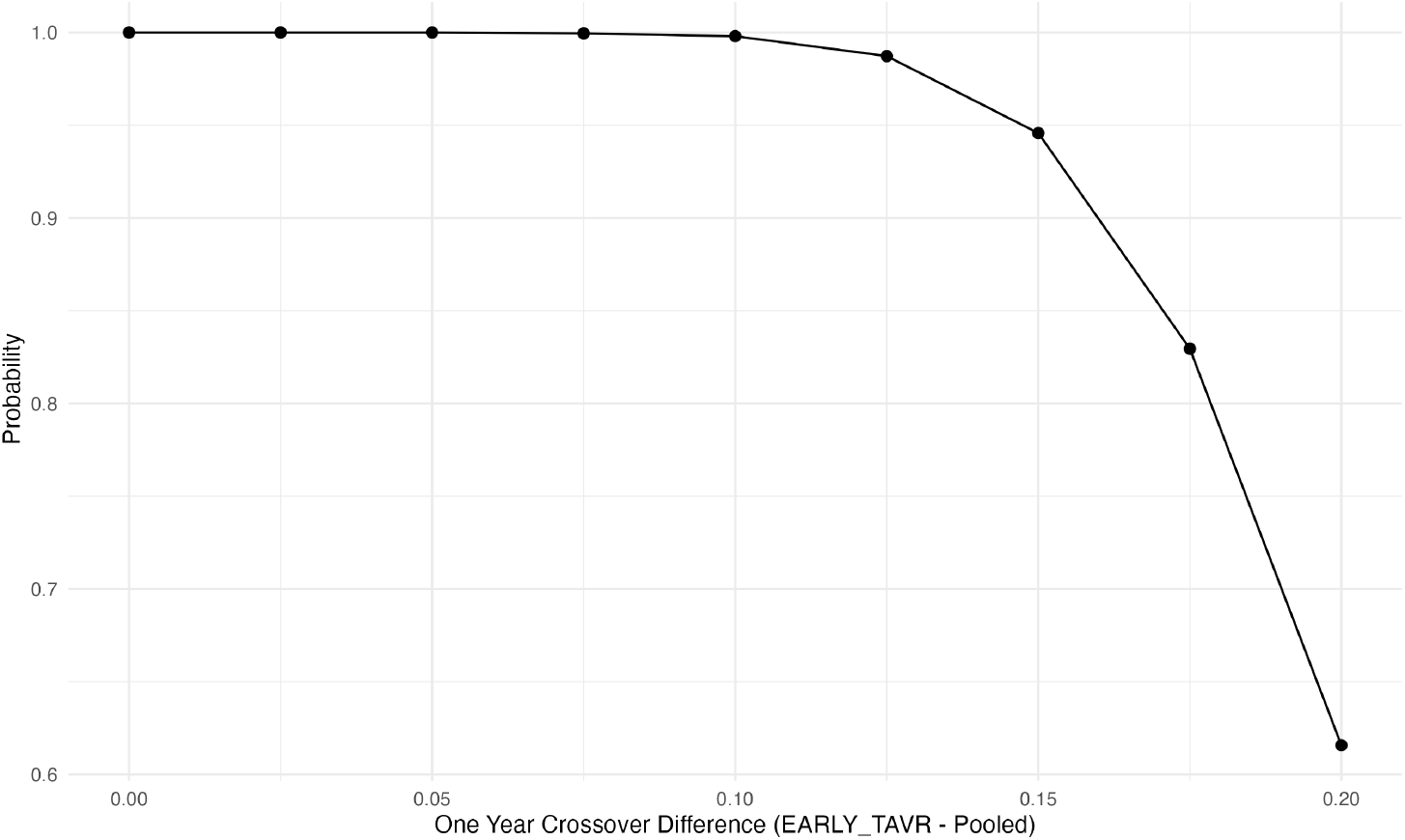
Probability of One Year Crossover Differences Probability of Differences Between EARLY _TAVR and Pooled Average of Other Trials

Given that subjects in all trials were asymptomatic at baseline, given the large heterogeneity in early crossovers between the single TAVR study and the other studies raising the possibility of an unblinding bias and given that any benefit from the intervention arm would be expected to be come more evident with time, a meta-analysis of the primary outcome was performed using one year landmarked data (Table 3 and Figure 4). Figure 4 shows that the pooled result from the three trials with the same approximate composite primary outcome (mortality and cardiac hospitalization) did not show any convincing evidence for a long term benefit of early AVR (HR 0.70, 95% CrI 0.34 - 1.08). Figure 4 also shows that the next study of long term outcomes with a one year landmark format is likely to produce a result that falls in an interval from 0.22 to 1.57. The Figure also shows the point estimates and confidence intervals of both the frequentist and Bayesian values for each individual trial. Due to the borrowing of information that accompanies a Bayesian (hierarchical) approach the individual intervals are shifted towards the global mean and slightly shrunken. As the Bayesian approach provides probability density functions, one can also examine not only the probability of a result of a ratio being greater or less than 1 but can also examine the probability of a clinically meaningful difference. For example, accepting a 10% or 20% reduction in the primary outcome as being clinically meaningful, the probability that the long term mean early AVR arm reduction reaches these thresholds is 85% and 70%, respectively. Finally there is a 19% probability that the long term outcomes from the next study could even give an unfavorable result for the early intervention arm (HR > 1).

**Figure 4.**
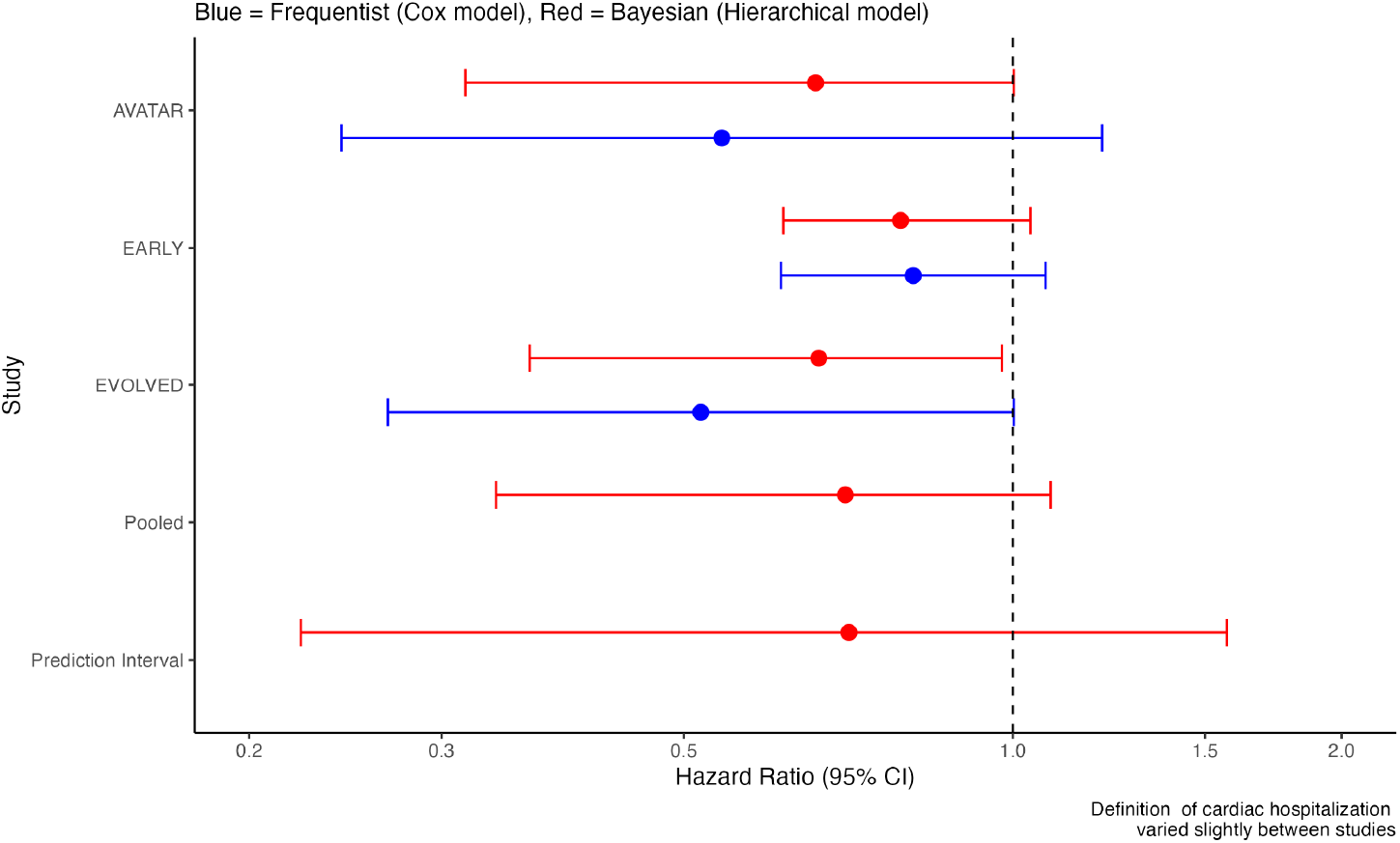
Bayesian Outcome Meta-analysis Forest Plot for Landmarked 1 Year Primary Outcome (Mortality, Stroke and Cardiac Hospitalization)

## Discussion

Four randomized trials investigating the early AVR treatment of asymptomatic AS compared to clinical surveillance were identified. In contrast to two previous meta-analyses[(7)](8) that concluded early AVR was associated with a significant reduction in the primary composite outcome of unplanned cardiovascular hospitalization, stroke or mortality, this meta-analysis suggests the long term benefit with early AVR for this outcome is uncertain (HR 0.70, 95% CrI 0.34 - 1.08).

As the same four studies have been considered, how to explain this discordance? Although the studies examined similar populations of asymptomatic severe AS individuals, as confirmed by exercise testing, important heterogeneity exists between the trials, especially in the choice of the active treatment intervention. The conclusions of the earlier meta-analyses[(7)](8) were driven by the EARLY_TAVR(3) study which accounts for 74% of the total outcomes (Table 2) and is the only trial to uniquely employ TAVR in the treatment arm. This trial is also the only trial with impressively high primary outcomes in the first year following randomization in the CS arm, driven principally by TAVR crossover hospitalizations. This early benefit contrasts with an absence in the surgical interventions trials and is hard to clinically rationalize as all subjects in the four trials were confirmed asymptomatic at the time of randomization with negative exercise testing. Why then did only EARLY_TAVR(3) patients became rapidly symptomatic with one year crossover rates approximately double that observed in the other trials. Also hard to understand is if TAVR truly reduces cardiac hospitalizations, why then does this benefit not continue to accrue after the first year? How to reconcile a landmark analysis that shows no such longer term benefit?

Given these observations and given also that the natural history of AS has likely not dramatically evolved in the decade since the first three trials were performed, it becomes necessary to seek other explanations for the large early TAVR benefit. What has altered in the past decade is the increasing availability of TAVR as a less invasive alternative to SAVR. Consequently our threshold to intervene has likely changed as “primum non nocere” in asymptomatic patients becomes less of an issue with the up front ease and safety of TAVR having miinimnal physical and emotional patient impact comapred to SAVR. It is relatively easy to see how a changing intervention threshold combined with the unblinded design of EARLY_TAVR(3) could have influenced the crossover rates and consequently cardiac hospitalizations. The decision to seek medical care and the decision for hospitalization both have subjective components and it has been previously documented that unblinded clinical trials with subjective outcomes have exxagerated effects sizes[RN54][RN55].

Specifically for EARLY_TAVR(3), there are 4 possible mechanisms that unbindedness, rather than TAVR effectiveness, could have contributed to the observed differences in crossovers and hospitalization rates. As all research participants sign an informed consent where the risks and benefits of trial participation are explained, the CS group knew they had severe disease and knew they were not being treated. This could increase anxiety and favor the conversion from an asymptomatic individual to a symptomatic patient — even in the absence of true progression of cardiac symptoms. Similarly the medical research staff, who are likely “believers” in the value of TAVR, knew which patients were not treated thereby increasing their probability of attributing any symptoms, whether of cardiac origin or not, to the underlying aortic stenosis. On the other hand, the early treatment group knew they were “fixed” and might be less likely to consult for any symptoms, again whether cardiac or not. The medical staff of the early TAVR patients knowing these patients had been treated might also be less inclined to attribute any symptoms to the cardiac disease. Importantly, all four of these possible scenarios would lead to a potential over-estimate of any differential hospitalization rates between early treatment and CS, implying a spurious early TAVR benefit.

The present analysis is limited by the unavailability of the origianl data as the individual trialists have refused data sharing. However, the ability to reasonably reproduce the published hazard ratios and cumulative incidence rates does provides some assurance that the early censored and landmark analyses reported herein are valid. On the other hand, a strength of this meta-analysis is its use of survival data which is generally preferred for meta-analyses of time-to-event outcomes as it allows for proper modeling of how risk changes over time. Previous meta-analyses[(7)](8) used aggregate data and were therefore unable to correct for a possible early benefit bias in EARLY_TAVR. The ability to perform time to event meta-analysis enables a bias corrected one year landmark analysis that adjusts for the time varying hazard ratios and thereby allows a more nuanced and powerful analysis for longer term outcomes.

Ultimately, the reader must decide whether EARLY_TAVR’s early benefits are due to a miraculous intervention or the normalcy of bias from an unblinded design. If TAVR truly decreases cardiac hospitalizations which scenario is more likely to be observed? A scenario with all the benefit occurring very early with no longer term benefits or a scenario where the benefit persists throughout the follow-up at a quasi constant rate or perhaps even slightly increasing with time as there vavle disease slowly progresses. If TAVR has a real impact, logic would dictate the later scenario is more likely. However the landmark analysis is compatible with former scenario, suggesting the observed difference is more likely due to bias from the unblinded design. This interpretation is also supported by EARLY_TAVR’s lack of benefit for the objective outcomes of death or stroke, past empirical evidence of bias in measuring subjective outcomes in unblinded trials[(14)](15) and the lack of a comparable early benefit with SAVR interventions. Overall, this supports the viewpoint that the EARLY_TAVR(3) benefit is more likley due to bias from unblinding than an actual benefit from early interventions in asymptomatic AS individuals.

Both previous meta-analyses[(7)](8) concluded, based on the Cochrane risk of bias tool(16), that these trials were all at low risk of bias. While this tool provides some general guidelines for bias evaluation, the tool can’t possibly codify all possible unique biases that may impact individual trials. Moreover the the Cochrane RoB tool provides no quantitative bias estimates. Based on this example, it would appear that simply checking boxes in the Cochrane RoB tool may be inadequate to exclude meaningful biases.

In conclusion, a detailed examination of the heterogeneity of the early outcomes between the four completed randomized trials of early intervention versus clinical surveillance in asymptomatic AS strongly suggests a potential bias due to unblindedness in the most recent and largest TAVR based trial. The unblinded nature of earlier trials was less problematic as the crossover required open heart surgery, a weighter decision than a TAVR procedure with a possible next day discharge. A proposed correction for this bias involving a one year landmark analysis shows no benefit for early AVR. The two published meta-analyses have not discussed this time varying heterogeneity or possible bias and their results are being used to justify viewpoints(2) calling for a modification of current guidelines(17) to abandon clinical surveillance for early active intervention as the treatment of choice for severe asymptomatic aortic stenosis. This manuscript suggests such a change would be premature and currently lacks convincing evidence.

## Data Availability

All data produced are available online at https://github.com/brophyj/asx_as

## References

1. Braunwald E. On the natural history of severe aortic stenosis. J Am Coll Cardiol [Internet]. 1990;15(5):1018–20. Available from: https://www.ncbi.nlm.nih.gov/pubmed/2312955

2. Lindman BR, Braunwald E, Pellikka PA. Aortic valve replacement for asymptomatic severe aortic stenosis-the time has come. JAMA Cardiol [Internet]. 2025; Available from: https://www.ncbi.nlm.nih.gov/pubmed/39937467

3. Genereux P, Schwartz A, Oldemeyer JB, Pibarot P, Cohen DJ, Blanke P, et al. Transcatheter aortic-valve replacement for asymptomatic severe aortic stenosis. N Engl J Med [Internet]. 2025;392(3):217–27. Available from: https://www.ncbi.nlm.nih.gov/pubmed/39466903

4. Loganath K, Craig NJ, Everett RJ, Bing R, Tsampasian V, Molek P, et al. Early intervention in patients with asymptomatic severe aortic stenosis and myocardial fibrosis: The EVOLVED randomized clinical trial. JAMA [Internet]. 2025;333(3):213–21. Available from: https://www.ncbi.nlm.nih.gov/pubmed/39466640

5. Kang DH, Park SJ, Lee SA, Lee S, Kim DH, Kim HK, et al. Early surgery or conservative care for asymptomatic aortic stenosis. N Engl J Med [Internet]. 2020;382(2):111–9. Available from: https://www.ncbi.nlm.nih.gov/pubmed/31733181

6. Banovic M, Putnik S, Penicka M, Doros G, Deja MA, Kockova R, et al. Aortic valve replacement versus conservative treatment in asymptomatic severe aortic stenosis: The AVATAR trial. Circulation [Internet]. 2022;145(9):648–58. Available from: https://www.ahajournals.org/doi/abs/10.1161/CIRCULATIONAHA.121.057639

7. Genereux P, Banovic M, Kang DH, Giustino G, Prendergast BD, Lindman BR, et al. Aortic valve replacement vs clinical surveillance in asymptomatic severe aortic stenosis: A systematic review and meta-analysis. J Am Coll Cardiol [Internet]. 2024; Available from: https://www.ncbi.nlm.nih.gov/pubmed/39641732

8. Brar SK, Leong DW, Razi RR, Moore N, Zadegan R, Mansukhani P, et al. Early aortic valve replacement in asymptomatic severe aortic stenosis: A meta-analysis of randomized controlled trials. Am J Cardiol [Internet]. 2025; Available from: https://www.ncbi.nlm.nih.gov/pubmed/40054514

9. Rohatgi A. WebPlotDigitizer. Available from: https://automeris.io

10. IntHout J, Ioannidis JP, Rovers MM, Goeman JJ. Plea for routinely presenting prediction intervals in meta-analysis. BMJ Open [Internet]. 2016;6(7):e010247. Available from: https://www.ncbi.nlm.nih.gov/pubmed/27406637

11. Riley RD, Higgins JP, Deeks JJ. Interpretation of random effects meta-analyses. BMJ [Internet]. 2011;342:d549. Available from: https://www.ncbi.nlm.nih.gov/pubmed/21310794

12. R Core Team. R: A language and environment for statistical computing [Internet]. Vienna, Austria: R Foundation for Statistical Computing; 2024. Available from: https://www.R-project.org/

13. Gabry J, Češnovar R, Johnson A, Bronder S. Cmdstanr: R interface to ‘CmdStan’ [Internet]. 2024. Available from: https://mc-stan.org/cmdstanr/

14. Page MJ, Higgins JP, Clayton G, Sterne JA, Hrobjartsson A, Savovic J. Empirical evidence of study design biases in randomized trials: Systematic review of meta-epidemiological studies. PLoS One [Internet]. 2016;11(7):e0159267. Available from: https://www.ncbi.nlm.nih.gov/pubmed/27398997

15. Wood L, Egger M, Gluud LL, Schulz KF, Juni P, Altman DG, et al. Empirical evidence of bias in treatment effect estimates in controlled trials with different interventions and outcomes: Meta-epidemiological study. BMJ [Internet]. 2008;336(7644):601–5. Available from: https://www.ncbi.nlm.nih.gov/pubmed/18316340

16. Sterne JAC, Savovic J, Page MJ, Elbers RG, Blencowe NS, Boutron I, et al. RoB 2: A revised tool for assessing risk of bias in randomised trials. BMJ [Internet]. 2019;366:l4898. Available from: https://www.ncbi.nlm.nih.gov/pubmed/31462531

17. Otto CM, Nishimura RA, Bonow RO, Carabello BA, Erwin JP, Gentile F, et al. 2020 ACC/AHA guideline for the management of patients with valvular heart disease: A report of the american college of cardiology/american heart association joint committee on clinical practice guidelines. Journal of the American College of Cardiology [Internet]. 2021;77(4):e25–197. Available from: https://www.sciencedirect.com/science/article/pii/S0735109720377962

